# Expert panel identified health-related social needs and methodological considerations for a polysocial risk score

**DOI:** 10.1101/2023.10.17.23297142

**Authors:** Joshua R Vest, Cassidy McNamee, Paul I Musey

## Abstract

**Objectives:** A polysocial risk score, which summaries multiple different health-related social needs (HRSNs) into a single likelihood of risk, could support more effective patient and population health management. Nevertheless, developing a polysocial risk score faces uncertainties and challenges due the HRSNs’ differing etiologies and interventions, cooccurrence, and variation in information availability.

**Methods:** A 3-round Delphi technique to elicit expert opinion and develop a preliminary polysocial risk score approach. Expert panel members included physicians (n=8), social service professionals & staff (n=9), and patients (n=6). For physicians, the primary qualification was direct patient care experience in screening or asking about patients’ health-related social needs. Social service professionals & staff had titles of: nurse, patient care assistant, patient advocate, community health worker, director of community services. Round 1 obtained an initial importance of HRSNs on general health & well-being and total healthcare cost. Panelists also suggested additional HRSNs Responses served as discussion points for Round 2. Five focus groups explored how HRSNs should be ranked; additional HRSNs to include; timing of measurements; management of non-response and missing data; and concerns about bias and equity. We analyzed the transcripts using a consensus coding approach. Panelists then completed a follow-up survey.

**Results:** Panelists identified 17 HRSNs relevant to health and well-being for inclusion in a polysocial risk score. Methodology concerns ranging from the sources and quality of data, non-random missing information, data timeliness, and the need for different risk scores by population. Panelist also raised concerns about potential bias and missaplication of a polysocial risk score.

**Conclusion:** A polysocial risk score is a potentially useful addition to the growing methodologies to better understand and address HRSNs. Nevertheless, development is potentially complicated and fraught with challenges.

## INTRODUCTION

Health-related social needs (HRSNs) encompass patients’ nonclinical, economic needs such as housing instability, food insecurity, and transportation barriers^1^. When unmet, these HRSNs create immediate risks to health^2^, increase utilization, and result in greater healthcare costs^3^. Moreover, the incorportation of social factor information has wide potential applicability in healthcare delivery. HRSN information can improve risk prediction models^4^, identify patients in need of social services^5,6^, and illuminate underlying disparities in population health^7^. As a result, HRSN screening has become more common among US healthcare organizations and will only likely become more important in light of the Centers for Medicare & Medicaid Services’ (CMS) new screening quality measures^8^.

However, translating information on patients’ HRSNs into action remains a challenge. For example, numerous HRSNs screening questionnaires exist, but most have no scoring guidelines^9^. HRSNs are often co-occurring^10^, but organizations often collect information on only select HRSNs^11–13^. Overall, HRSNs are not view holistically. Likewise, HRSN information is often stored in different locations within EHRs, thereby inhibiting a comprehensive view of the patient^14^. Most importantly, despite increased screening, most patients’ HRSN go unmeasured and unresolved^15^.

A polysocial risk score, which summaries multiple different HRSNs into a single likelihood of overall risk, could be a method that supports more effective use of patients’ social factor information^16,17^. This idea of summarizing risk into a single measure has analogs in genetics’ polygenic risk scores, clinical care’s various comorbidity indices, or public health’s area-level community vulnerability indices. Additionally, limited applications of an overall social risk score concept are suggestive of its potential usefulness. For example, counts of HRSNs have been associated with diabetes^18^, cancer^19^ and older adult mortality in US national surveys^20^ as well as cardiovascular disease in national Korean survey data^21^.

Nevertheless, developing a polysocial risk score faces uncertainties and challenges. HRSNs have differing etiologies and require different interventions^22^. Also, the examples of previous social risk scores noted above that simply count the presence of different risk fail to account for health-related social needs’ interrelated nature and the need to prioritize some factors over others during clinical care delivery^23^. Importantly, relevant HRSNs data may go uncollected or patients may not supply all relevant information^24,25^.

We sought to lay the foundations for the development of a polysocial risk score appropriate for the general US adult patient population. To address the above uncertainties and challenges, we leveraged a nationwide expert panel to identify relevant HRSN, to assess relative importance of each HRSN, and to provide methodological guidance. Specifically, expert panels can identify which factors should be included in risk scores, ratings can be used to guide the relative weighting of factors in a score^26^, and experts can identify approaches to account for missing information^27^. Our work sets the stage for the future development and quantitative assessment of a polysocial risk score.

## METHODS

We used a 3-round Delphi technique^28,29^ to elicit expert opinion and develop a preliminary polysocial risk score approach. Our mixed-method approach used focus groups and two rounds of surveys.

### Expert Panel Formation

We recruited 23 individuals to participate in our expert panel (2 individuals declined our invitation) representative of physicians (n=8), social service professionals & staff (n=9), and patients (n=6). For physicians, the primary qualification was direct patient care experience in screening or asking about patients’ health-related social needs. Following our previously successful approach^30^, we emailed authors of research studies on HRSNs or the role of HRSNs in medical education and practice. We were attentive to geographical diversity (CA, DC, FL, IL, IN, MA, MD, NM, NY, OH, PA) and practice setting (e.g., emergency department and primary care). To identify panelist representing social services, we requested each recruited physician to recommend one case manager, social worker, or other relevant staff (e.g., nurse, patient care assistant, patient advocate, community health worker, director of community services) within their institution to participate. We also relied on our professional networks. Patients were recruited through existing contacts with health systems. Panelists self-reported gender, age, and race/ethnicity to ensure diversity (see Table 1). We offered an incentive to all panelists for participation. The literature suggests between 10 to 18 participants for Delphi panels^31,32^, because we sought the inputs of multiple different disciplines and patients, we exceeded this recommended panel size.

**Table 1.**
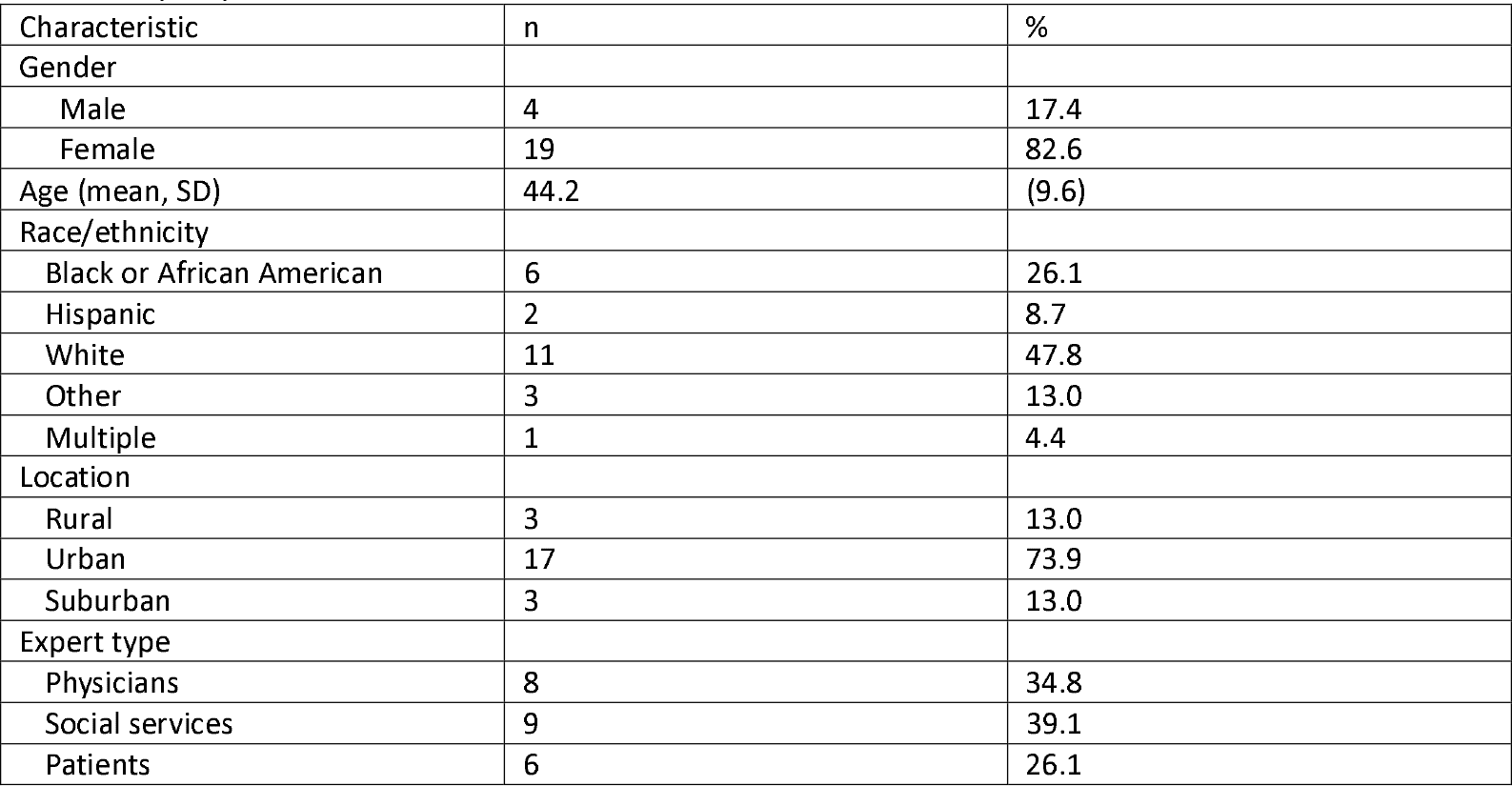
Expert panel member demographics.

| Characteristic | n | % |
| --- | --- | --- |
| Gender |  |  |
| Male | 4 | 17.4 |
| Female | 19 | 82.6 |
| Age (mean, SD) | 44.2 | (9.6) |
| Race/ethnicity |  |  |
| Black or African American | 6 | 26.1 |
| Hispanic | 2 | 8.7 |
| White | 11 | 47.8 |
| Other | 3 | 13.0 |
| Multiple | 1 | 4.4 |
| Location |  |  |
| Rural | 3 | 13.0 |
| Urban | 17 | 73.9 |
| Suburban | 3 | 13.0 |
| Expert type |  |  |
| Physicians | 8 | 34.8 |
| Social services | 9 | 39.1 |
| Patients | 6 | 26.1 |

### Round 1 – Preliminary importance, ranking, and factor identification (survey)

Round 1 obtained an initial importance rank order of each selected HRSN on the outcomes of general health & well-being and total healthcare cost. Risk score development requires a clear definition of the outcome of interest^27^ and these outcomes are influenced by HRSNs. Outcomes are relevant to patients, healthcare organizations, and policy makers. Panelists rated the importance of housing instability, financial strain, food insecurity, legal problems, transportation barriers, unemployment and social isolation (definitions provided) on a Likert-type scale ranging from *absolutely critical to not important at all* for both study outcomes. Panelists then ranked the same HRSNs from *most important* to *least important* for the two outcomes. We selected these HRSNs as a starting point as these domains are frequently represented in screening questionnaires. Panelists were also able to suggest additional HRSNs for consideration and provide their assessment of importance. The survey was administered in REDCap^33,34^ (see Appendix). We analyzed the responses using means and stratified to identify potential differences. Suggestions were compiled into single lists by outcome. These findings were used as discussion points and to stimulate conversation during the Round 2 focus group data collection described below (see Appendix A1 & A2).

### Round 2 – Focus groups

We conducted 2 physician (MD), 2 social service professionals & staff (SO), and 1 patient (PT) focus groups with shared digital whiteboards. Prior to each focus group, we reviewed the objective of the study and shared the Round 1 survey results. JV moderated each focus group and covered the following topics using a semi-structured interview guide:

1. *Importance and ranking*. For each outcome, we reviewed the results and discussed which factors should be considered of higher weight within a polysocial risk score.
2. *Consideration of additional factors*. Discussion reviewed the additional factors suggested by the panelists and specifically sought views on the inclusion of behavioral factors like physical activity and nutrition.
3. *Measurement timing*. We sought opinions on the timing of measures relevant for the risk score, such as within the past 30 days, past 3 months, past 6 months, or past year.
4. *Handling of missing data*. This area of discussion elicited guidance on how non-response and missing data should be managed in the polysocial risk score.
5. *Potential concerns*. We also asked about specific concerns around the use of a polysocial risk score in particular with respect to bias and equity.

Each focus group lasted an average of 93 minutes and was recorded and transcribed with consent. After each focus group, we met to review notes and to assess saturation.

We analyzed the transcripts using a consensus coding approach^35^. Three team members read one transcript independently and created codes using an open coding approach. Through a joint reading session, coding was compared and refined to develop an initial code book. Pairs of coders read and coded the remaining transcripts using this codebook independently and then resolved differences through joint readings. When consensus could not be obtained, the third member of the team served as the final adjudicator. We summarized codes into overall themes and identified representative quotes.

### Round 3 – Final importance and ranking (survey)

After the completion of focus groups, all panelists completed a follow-up survey using 5-point Likert-type items to assess 17 different HRSNs’ (from the round 1 survey and those identified by focus groups) risk to health from very low to very high (see Appendix A3 & A4). We aligned the suggested factors to the domains identified by the IOM for consistent language, when possible^36^. Based on feedback obtained during the focus groups, we did not include questions specific to healthcare costs. To better understand potential usage of a polysocial risk score, we also asked panelist to rate the potential effectiveness of the polysocial risk score in each of the five health system social care activities identified by the National Academies of Medicine: awareness, adjustment, assistance, alignment, and advocacy^37^. Rankings were on a 5-point scale and definitions of each activity were provided. We also added a single global question to assess the appropriate level for using a polysocial risk score: individual-level care, population-level activities, or both. We calculated medians stratified by participant type^38^ and compared using the Kruskal-Wallis test.

The study was approved by the Indiana University IRB (#17119).

## RESULTS

Through a combination of qualitative and quantitative data, the expert panel provided guidance on why particular HRSNs should be included in a polysocial risk score, methodological considerations, and the positives and potential negatives of the usage of such a score (Table 2).

**Table 2.** Overarching themes and codes from expert panelists’ views on a polysocial risk score.

| Theme | Code | Description | Representative quote |
| --- | --- | --- | --- |
| Social factor domains |  | Explanation or justification for which HRSNs (domains) would be more relevant or viewed as more important |  |
|  | Available measurement | Screening tool or questionnaire exists (whether or used or not) to measure the factor. | "The violence measures...Yes, they're poor measures, but they are a measure that is recorded, however. They might be at each institution, so it's something that you could easily, potentially pull..." (MD#3) |
|  | Available interventions | Resources, programs, interventions, exist or are at least in theory available to address the issue. | "I wonder how much that points out our focus as providers on things we can fix." (MD#4) |
|  | Personal experience | Personal experiences or stories of how the factor was important in life and care. | "Growing up on a in a rural area, transportation was a huge issue." (PT#3) |
|  | Relevance to health | Panelist thought highly important or strong determinant of health that justified its inclusion. (explicitly stated) | "Education...probably the most strongly correlated thing, for many reasons, with health for all sorts of outcomes." (MD#7) |
|  | Role of behavioral factors | Thought / opinion on specifically including behaviors (nutrition, physical activity, smoking, etc.) in the single risk score. | "I would personally separate them, just because I think that the interventions are different...We focused a lot of attention on behavioral modification for like a hundred years and then realize that it wasn't enough. And that's why there's been kind of this pivot to social risk or social need." (MD#5) |
| Risk score methodology |  | Descriptions of "how" a polysocial risk score could be specified in terms of inputs, transformations, and analyses. |  |
|  | Data sources for risk score | Which data sources, structured, unstructured, surveys would be appropriate or inappropriate for inputs into the score. | "I think it definitely needs to be pulled from a number of different sources. I think a survey alone doesn't really provide the response or the feedback that we can get that someone that a nurse wrote as a note could give." (SO#9) |
|  | Handling missingness in data | Approaches, methodologies, alternatives or concerns for HRSNs that are unavailable due to patient nonresponse, never asked, new patients, no screening tools, willingness to share. | "A.I. might be able to help, even though I'm not a terrible fan. Real time algorithmic imputing of data based on what they do respond. We'll navigate the false positives probably. But if you've got the resources to actually address the needs that should come or become evident afterwards." (MD#3) |
|  | Timeliness | How recent must data be to be useful in risk scores. | "Needs kind of go up and down...If you're using [risk score] for payment, adjusting payment, then you may want to capture a much wider timeframe. If you're using it for clinical care, you may want to focus on current. So there's no one answer. It really depends on the on the use case." (MD#6) |
|  | Generalizability vs stratification | Assessments of whether a risk score is broadly applicable to all patients or if different scores need developed for different populations (e.g., stratification, contextualized, tailored). | "The rankings may be different, even if we're thinking, like geographically, the impact of a transportation barrier. If you're someone who's living in a rural neighborhood versus an urban neighborhood, I'm gonna weigh those very differently." (SO#2) |
| Application |  | Purpose and consequences of a polysocial risk score |  |
|  | Concerns for bias & stigma | Perceptions of potential inequities, disadvantage, or systematic errors in the risk score or any potential negative effects from its application. | "So they're gathering information. They're gathering your data. They know how much you make. They know where you live. They know you know how much healthcare, how much you're using, how many visits you have so it would not surprise me that this is what they would be used for, but it also scares me in sense that they could then take this information and start charging poor people more." (PT#3) |
|  | Usage | Appropriate and inappropriate applications of the risk score in clinical care or population health management activities. | "You could probably use a score to provide a threshold of when to refer to an intervention." (MD#1) |

### Which factors should be included in a polysocial risk score: reasons for inclusion

The panel identified 5 broad reasons for inclusion of specific HRSNs in a polysocial risk score: *relevance to health, available measurement, available interventions*, and *personal experience*. These same reasons also explained which factors were viewed as more important or more critical for inclusion than others. In general, all the HRSNs discussed by the expert panel were recognized as important determinants of health and well-being. A physician summarized it thusly: “*They’re all critically important to health…and everything is interconnected* (MD#5).” Nevertheless, several needs were identified as more important due to their explicit *relevance to health*. For example, a director of community services described issues of financial strain, food insecurity, and housing instability as having “*a little bit more urgency* (SO#2)”, a social worker called them “*root causes* (SO#1)”, and a patient equated those same factors as reflective of Maslow’s hierarchy of needs (PT#1). A physician identified a potential pathway for these same issues to affect health: “*Cost of daily living, buying food, paying your rent, keeping the utilities active in your home, and many people, especially those who have children, are going to prioritize the family’s stability over their own personal health* (MD#8).” At the same time, panelists noted that lower ranked, or even other factors, were critically important to health. Such examples included: exposure to violence, legal problems, health literacy, discrimination, education, and adverse childhood experiences.

Relatedly, several factors were noted as more relevant than others due to *personal experience*. While some providers and staff recalled care delivery events, patients more frequently described particularly salient events or contexts that warranted particularly including some health-related social needs. For example, a patient introduced the immigration status: “*I go to church every Sunday, and I go to a Spanish branch and most of them, and if not all of them, have legal documents problems. And so I just see them struggling more* (PT#5).” In similar fashion, patients from rural areas (PT#3 & PT#6) recounted how transportation barriers posed a significant barrier to their health.

Beyond general importance, providers and staff often noted the ability to measure (*available measurement*) and intervene (*available interventions*) as the logic for inclusion and greater importance of some HRSNs over others. For example, one physician noted: “*We may tend to rank these things with things that we can do something about right? So food insecurity is one where people find it a little bit easier to refer people to the 8 million programs that are out there versus solving a senior citizen social isolation, which is way harder* (MD#1).” Another physician illustrated the concept through a contrast. They stated: “*Certainly they’re all associated with health outcomes. But food and security*…*in terms of operations it’s something that a lot of people are thinking about clinically and…there are state programs and local programs. Social isolation - I think a lot of people shy away from this, because it maybe seems less tangible to address it in clinical settings* (MD#7).” Another physician panelist made a similar contrast, but with financial strain, unemployment, and social isolation: “[Financial strain and unemployment] *are measurable and potentially addressable by interventions that we can think about providing in a way, that to me, social isolation might be more challenging to get at* (MD#4).”

Conversely, the general view of panelists was to exclude behavioral factors (e.g. nutrition, physical activity, etc.) from consideration of a polysocial risk score. The justification offered by panelists was generally that “*the interventions are different* (MD#5)”, the “*solution for those two areas is different* (SO#4)”, and HRSNs and behavioral factors require “*totally different resources* (SO#1).”

The final survey rankings (Round 3) for the identified HRSNs also reflected the general perception that many different HRSNs were important to overall health & well-being (Table 3); specifically, on a 5-point Likert scale, every social factor had at least a median rating of 3. The highest median score was for access to care. The next highest rating factors mirrored much of the discussion and included: adverse childhood events (ACEs), discrimination, exposure to violence, financial strain, food insecurity, housing instability, language barrier, and stress. The only social factor where rating varied significantly (p=0.0107) was immigration status, where patients had a higher median rating.

**Table 3.** Expert panel’s second round median survey rating of suggested health-related social needs’ risk to health & well-being from very low risk to health (1) to very high risk to health (5).

| Social factor | Overall | Physicians | Social services | Patients | p |
| --- | --- | --- | --- | --- | --- |
| Access to care | 5 | 5 | 4.5 | 5 | 0.1187 |
| Adverse childhood events (ACEs) | 4 | 4 | 3.5 | 4 | 0.9159 |
| Discrimination | 4 | 3.5 | 4 | 3 | 0.0806 |
| Exposure to violence | 4 | 4 | 4 | 4 | 1.000 |
| Financial strain | 4 | 4 | 4 | 5 | 0.1906 |
| Food insecurity | 4 | 4 | 5 | 4 | 0.2286 |
| Housing instability | 4 | 4 | 4 | 4 | 0.9768 |
| Language barrier | 4 | 3.5 | 3.5 | 4 | 0.7420 |
| Stress | 4 | 3.5 | 4 | 4 | 0.1727 |
| Childcare availability | 3 | 3 | 3 | 3 | 0.9327 |
| Education level | 3 | 4 | 3 | 3 | 0.1877 |
| Health literacy | 3 | 3.5 | 3 | 4 | 0.8342 |
| Immigration status | 3 | 3 | 3 | 4 | 0.0107 |
| Legal problems | 3 | 3 | 2 | 3 | 0.2828 |
| Social isolation / loneliness | 3 | 3.5 | 4 | 3 | 0.3297 |
| Transportation barriers | 3 | 3 | 4 | 3 | 0.3993 |
| Unemployment | 3 | 3 | 3 | 3 | 0.5164 |

### How could a polysocial risk score be constructed: risk score methodology

Panelists’ descriptions of “how” a polysocial risk score could be constructed covered several concepts related to the necessary inputs, data transformations, and analyses. First, panelists were very broad in their considerations of the data sources and types that could be used within a polysocial risk (*Data sources for risk score*): surveys, structured EHR data, unstructured notes, and diagnosis codes. Part of the logic was any one source could be insufficient. For example, one social worker noted: “*I think a survey alone doesn’t really provide the response or the feedback that we can get that someone that a nurse wrote as a note could give. We need to be pulling information from all places* (SO#9).” In terms of specific views, survey data was viewed as “*cleaner* (SO#5)” and viewed very favorably by panelists working with social service organizations, because it was “*patient reported* (SO#1, SO#2 & SO#4)”. At the same time, panelists were aware that data sources had limitations. Concerning ICD-10 Z codes, one physician observed: “*You never know when it was documented. But I think relying on them would be certainly underestimating a lot* (MD#7)”. Another nurse echoed this sentiment: “*I don’t know any consistent provider who actually uses those* [Z] *codes…It’s not gonna be like super robust* (SO#8).” Similarly, a social worker noted: “*I do think there are other elements that absolutely can be pulled from the record you just need to be thoughtful about what exactly. For example: problem lists. Problem lists are notoriously inaccurate* (SO#1)”.

While surveys had advantages, a population health director noted their organization’s observed challenges with missing information in screening surveys: “*The full completion rate is about 15%, which means they’re leaving a lot blank* (SO#5).” Echoing the potential for missing information, a patient noted: “*So everything the computer got is because the patient somehow shared*.” (PT#5) Multiple physicians (MD#1, MD#3 & MD#4) reported that missing information was not at random. Options for handling missing information ranged from statistical imputation (MD#1, MD#3, MD#8 & SO#1) to re-screening (“*we are screening multiple times, so we hope that they would answer eventually* (SO#1)”). The role of imputation was generally caveated to be limited to usage of the polysocial risk score for research purposes. This restriction was in the context of the recognition that “*prefer not to answer and I just skip that question* (MD#6)” were fundamentally different. A patient panelist confirmed this view: “*Somebody may miss a question because they don’t understand it. They prefer to leave it blank than give inaccurate information* (PT#2)”. Reflecting this potential, a social worker reported that their organization followed up with all individuals who declined screening to still offer resources (SO#3).

A physician panelist captured the tensions in the discussions of *timeliness* (how recent must data be to be useful in risk scores) well: “*I mean ideally, that time of visit, right? That’s when you’re making decisions. But that’s not practical* (MD#3).” In favor of very recent information, a social worker stated: “*I want to know where my patient is today* (SO#7).” Likewise, panelists noted that social program benefits can change frequently and that some issues like homelessness require immediate attention (SO#3). Similarly, a patient panelist endorsed the value of more recent information when s/he said, *“It depends on what, what day it is as to which one of these things are most important* (PT#6).” Nevertheless, panelists noted the burden of “*survey fatigue* (MD#1)” for patients, the “*risk of losing trust* (SO#1)” if screening was not tied to available resources, and that for some percentage of patients HRSNs can remain “*unchanged* (MD#4)” for months. Longer time frames were still seen as potentially useful. A physician noted, “*even having that sense of the past in there can give you some sort of data that is relevant to like health outcomes* (MD#7).” Likewise, longtime frames were *“easier to define like a year of observation for so many reasons, including insurance and eligibility* (MD#2). Choices of specific timing were attributed to “*kind of a pragmatic decision* (MD#5),” trying to strike “*a good clinical balance as well as a good logistic balance* (MD#1),” or what was defined by the organizations’ screening tools (SO#3 & SO#9). Overall, a social worker summarized: “*As a clinician, a history is important for me regarding treatment outcomes. But in terms of an accurate current risk score, a more recent timeframe is more helpful to me* (SO#6).”

Panelists noted important ways that a polysocial risk score may lack *generalizability* or need *stratification*. For example, immediate needs, and the ability to address patient needs, vary between emergency department and primary care settings (MD#8), patients may experience very different “*built economic and social environments* (SO#4),” and older patients face different financial risks than younger patients (MD#4). Patient panelists were particularly attentive to the differences between rural and urban populations and the need to consider those different contexts within a polysocial risk score (PT#3, PT#5 & PT#6).

### What can be done with the polysocial risk score: application

The broader theme of *application* of a polysocial risk score included *usage* and *concerns for bias and stigma*. In terms of *usage*, the overall responses suggested that a polysocial risk score would be “very effective” for all 5 social care activities suggested by the National Academies of Medicine (Table 4). However, these general views differed by panelist type. Physician panelists reported significantly lower perceptions of effectiveness for adjustment (p=0.0144), alignment (p=0.0202), and advocacy activities (p=0.0312). A physician panelist noted: “*I think you actually need individual items largely for adjustment. If you’re gonna do telehealth follow up, you need very detailed information about like digital access or transportation…That does not negate the value for other activities for other use cases* (MD#6).” Another had a similar concern, *“I worry about the creation of a single unique score because it kind of disaggregates our ability to act on the information. If you tell me the risk score is 13. I can’t do anything. Rather telling me that they screen positive for housing and food insecurity and something else*…*I might have an intervention to deliver, or at least resources to provide* (MD#4).”

**Table 4.** Expert panel’s second round survey ranking of potential effectiveness of a polysocial risk score (not at all effective (1) to extremely effective (5)) for social care activities.

| Social factor | Overall | Physicians | Social services | Patients | p |
| --- | --- | --- | --- | --- | --- |
| Awareness | 4 | 4 | 4 | 4 | 0.6305 |
| Adjustment | 4 | 2 | 4 | 4 | 0.0144 |
| Assistance | 4 | 2.5 | 4 | 4 | 0.2679 |
| Alignment | 3 | 2 | 4 | 3 | 0.0202 |
| Advocacy | 4 | 2.5 | 4 | 4 | 0.0312 |

In terms of the level for which a polysocial risk score would be most suited, physicians skewed to more population-level (62.5%) usage (Figure 1). The remainder of experts were more broadly distributed. In discussions, panelists speculated about the role of a polysocial risk score at a population level for stratification purposes. For example, a physician noted, “*Population level stratification I still think that it is a starting point for the care teams to decide. Which subpopulation of patient I should start with questioning, or which one requires more frequent or in-depth assessment of their social needs, and then based on those specific needs - I take action* (MD#2).” Other physicians used phrases like “*distinguish which track of intervention patients should go in* (MD#5)”, to “*triage* (MD#7)” patients, or for “*trying to find people* (MD#1).” Social service professionals & staff also reported similar applications at the population level. For example, a health equity program director observed: “*I think it is helpful…aggregating data, so that you can start to see patterns or not see patterns. That’s really valuable for allocating resources* (SO#1).” Likewise, a director of community services mentioned such a score might support “*a better population-level understanding of what’s happening in our institutions* (SO#2).”

**Figure 1.**
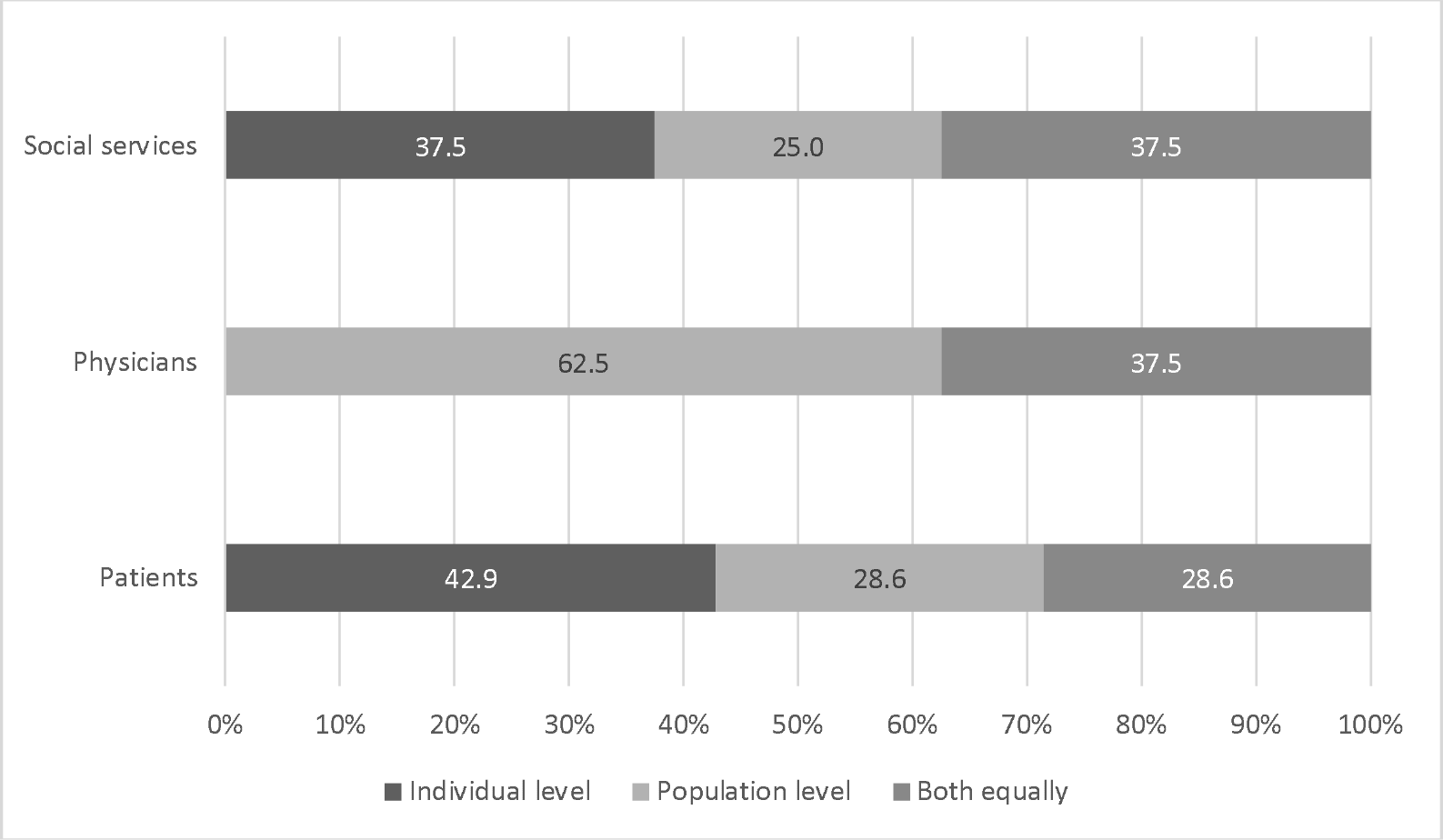
Expert panelists’ views on the level of activity for which a polysocial risk score is most suited.

Panelists of all types noted the potential for bias and stigma both specific to the idea of a polysocial risk score, but also for social factor data collection in general. In terms of potential biases in the data from nonresponse, a patient reported “*I don’t want to get myself in trouble…So I think there can be bias or answers that are not accurate answers* (PT#5).” Another patient agreed, “*Everything’s not fair game…Something from 10 years ago, it’s not really relevant anymore* (PT#6).” Another said simply “*If they don’t need to know, they don’t need to know* (PT#2).” Biases in the data could also arise from structural and systemic problems in healthcare access and society. A physician noted: “*If you condition or train your algorithm on costs, you actually like bake in racial disparities because the cost of healthcare and the cost of spending we tend to spend more money on folks who are white* (MD#4).” A physician noted: “*Some people don’t have access to care…,the implicit bias in the healthcare system, and the patterns of cost of care is very different* (MD#2).”

These biases could translate into a problematic application of a polysocial risk score. A director of community services asked: “*How much of these scores are really just going to reflect more of that and how poorly resourced our system and our structured system is, and we’re just continuing to reinforce that…*(SO#2).”Likewise, a physician summarized the concerns as: “*privacy and being used for some communities more than others, the potential* [for] *being reductionist and sort of leading to disparities, and how patients are treated* (MD#7).” However, even with these concerns, some panelists were in favor of a single polysocial risk score. For example, a physician noted parallels to genetic risk scores: “*The concern for discrimination is accurate*…*Unfortunately, it’s gonna happen anyways*…*I think you have to be very careful on how you present this…I don’t think that’s necessarily a hard stop either* (MD#3).” Another physician stated: ““*I’m a person that believes in doing a score, mostly to trigger interventions to do things… I absolutely worry about bias. increased child protection reports…differential interactions…differential treatments* (MD#1).” Similarly, another stated “*I can see a potential benefit of a single score…I think that there are a lot of doctors out there who want to help and who want to do good things with the data. There are a lot of people out there who might abuse the data and use it to transmit stigma*…(MD#5).”

## DISCUSSION

Our expert panel identified a set of HRSNs relevant to health and well-being for inclusion in a polysocial risk score. However, the set of recommended HRSNs tended to be broader than what healthcare organizations currently collect^12,13^. This may be resolved given CMS’ increased screening requirements or if future reimbursement policies include Z-codes. However, the potential for the expert panelists’ ratings to inform a weighting approach for a polysocial risk score was less clear as tended to rate all these factors highly in terms of risks to health. It may not be possible to assume any differential weighting of HRSNs beyond some very broad categories. Alternatively, the high rating of all factors may indicate that the existing approaches of simply counting unweighted factors may be most appropriate^19,20,39^. Yet still, given that the panelists tended to see all factors as relevant and interrelated, empirical methods may be most appropriate for establishing weights.

Methodology concerns ranged from the sources and quality of data, non-random missing information, data timeliness, and the need for different risk scores by population. These challenges are generally the same ones faced by any risk score methodology^27^. However, the nature of HRSN data may exacerbate some of these challenges. For example, HRSNs are known to change over time^40–42^, which is different from polygenic risk scores or a comorbidity index. Additionally, no agreed-upon measurement frequency really exists. Further, while clinical information systems like EHRs include data on many HRSNs^30^, these systems/processes of care delivery were not designed to systematically collect HRSN data. Therefore, any effort to measure the HRSNs needed for a risk score will require multiple modalities, e.g., surveys, natural language processing, or inclusion of social service referral history.

Based on the identified challenges and the opinions of the expert panel, the clearest use case for a polysocial risk score may be in increasing awareness, that is identifying those with HRSNs^37^. However, it is possible awareness could be best suited to the population-level rather than individual care. Healthcare organizations, physicians, and patients realize actions must follow identification of HRSNs^43^. The aggregated, summative nature of a risk score combined with challenges in measurement timing, could conflict with physicians’ and staffs’ need to know specific individual-level HRSNs and pressing needs in order to adjust care or to provide assistance directly or through referrals. Instead, population-level awareness would be conceptually closer to risk stratification^44,45^ where organizations would be able to identify larger patient groups for additional screening or for follow-up activities^46^. Stratification based on polysocial risk score s could be a means to more effectively use resources for organizations that are responsible for screening large numbers of patients^15^. Additionally, a polysocial risk score could be useful as an adjustment factor in quality reporting metrics^47^.

Regardless, bias in inferences about patients and misuse of the score are paramount concerns to the expert panel as summary risk scores for numerous conditions and risks have been demonstrated to be biased^48,49^. Selective reporting and collecting of HRSNs creates distorted picture of risk^50^. Moreover, HRSN data are particularly susceptible to biases due to differential access to care, explicit, and implicit biases^51^. Underserved and underrepresented populations are disproportionally burdened by HRSNs^52^ heightening the need to ensure a polysocial risk score does not have negative and differential consequences on care. Of course, all of these concerns about a polysocial risk score hold true for individual HRSN data collection.

### Limitations

The study excluded behavioral factors, which others have combined with HRSNs^53^. We may have seen different ratings if we had included behavorial factors. While we had a diverse set of physicians, patients, and social service staff, and observed saturation in our focus groups, it is possible a different set of experts may have generated different preferences and opinions. In particular, expansion of the panel to executive health system leadership, policy makers, or payer representatives may have identified additional themes and HRSNs.

## CONCLUSION

A polysocial risk score is a potentially useful addition to the growing methodologies to better understand and address HRSNs. Nevertheless, development is potentially complicated and fraught with challenges.

## Data Availability

Data are not available.

## Funding

This work was supported by the Health Disparity and Equity Research Program at the Regenstrief Institute, Indianapolis, IN.

## Acknowledgements

The authors thank the panelists and Ms. Brittany Miller for assistance in coding.

## Notes

### Competing Interest Statement

Joshua Vest is a founder and equity holder in Uppstroms, LLC a technology company.

### Author Declarations

The IRB of Indiana University IRB gave ethical approval for this work (#17119).

