## Appendix for "Expert panel identified health-related social needs and methodological considerations for a polysocial risk score"

A1. Round 1 health-related social needs rating, ranking and suggestion survey items.

1. Please rate the importance of each of the following social risk factors for an adult patient's overall general health and well-being from absolutely critical to not important at all. (see A2 for items).
2. Are there other social risk factors, not listed above, that are important for an adult patient's overall general health and well-being?
3. Based on your opinion, please rank the following social risk factors from most important to least important for an adult patient's overall general health and well-being. (see A2 for items).
4. Please rate the importance of each of the following social risk factors for an adult patient's overall healthcare costs from absolutely critical to not important at all. (see A2 for items).
5. Are there other social risk factors, not listed above that are important for an adult patient's overall healthcare costs?
6. Based on your opinion, please rank the following social risk factors from most important to least important for an adult patient's overall healthcare costs. (see A2 for items).

A2. Expert panel’s first-round mean rankings of social factor importance.

|  | General health | Health care costs |
| --- | --- | --- |
| Housing instability | 2.0 | 2.6 |
| Financial strain | 2.2 | 2.0 |
| Food insecurity | 2.8 | 4.0 |
| Unemployment | 5.0 | 4.5 |
| Social isolation | 5.1 | 5.1 |
| Legal problems | 5.2 | 5.1 |
| Transportation barriers | 5.7 | 4.5 |

^1^Lower score is more important.

A3. Social factors and definitions included in Round 2 survey.

| Social factor | Definition provided to expert panel |
| --- | --- |
| Housing instability | housing disruptions or related problems, from frequent moves or difficulty paying rent to being evicted or being homeless^1^ |
| Financial strain | the perceived ability of one’s income to fulfil financial obligations;^2^ state of being wherein a person cannot fully meet current and ongoing financial obligations, cannot feel secure in their financial future, and is unable to make choices that allow them to enjoy life. Opposite of “well-being”^3^ |
| Food insecurity | household lacks access to adequate food because of limited money or other resources^4^ |
| Legal problems | criminal justice matters or events involving arrest and appearance in court to answer a charge^5^ |
| Transportation barriers | access to modes of transportation and burdens by distance / travel time^6^ |
| Unemployment – | not working, but wants to work; excludes those voluntarily / involuntarily out of workforce (full-time students, homemakers, retirees, and the disabled)^7^ |
| Adverse childhood events (ACEs) | potentially traumatic events that occur in childhood (0-17 years)^8,9^ |
| Stress | a subjective state arising when an individual believes that he or she does not possess the resources to cope with a threatening situation, resulting in tension, restlessness, nervousness, or anxiousness^8^ |
| Health literacy | the degree to which individuals have the capacity to obtain, process, and understand basic health-related decisions^8,10^ |
| Discrimination | unequal treatment based on physical characteristics or social group assignment^11^ |
| Language barrier | limited English language proficiency^12^ |
| Education level | number of years of time formally spent in school and the highest degree earned^8^ |
| Access to care | timely use of personal health services to achieve the best possible health outcomes (i.e. effective and realized access)^13^ |
| Immigration status | – a person who came to the United States as an adult, regardless of actual legal status^15^ |
| Exposure to violence | actual or threatened physical, sexual, psychological, or emotional abuse by a family member, caregiver, current or former spouse or partner, or dating partner^8^ |
| Childcare availability | child care services that meet both the child's needs and the parent's work schedule^14^ |
| Social isolation / loneliness | feeling that intimate and social needs are not adequately met^15^ |

A4. Round 3 health-related social needs rating, ranking and suggestion survey items.

1. Please rate the following factors in terms of risk to patients' overall health and well-being. (see A3 for items).
2. For each of the use cases below, please rate potential effectiveness of a polysocial risk score:

| identifying the social risks and assets of defined not at all effective patients and populations (awareness) | not at all effective  slightly effective  somewhat effective  very effective  extremely effective |
| --- | --- |
| altering clinical care to accommodate identified social barriers (adjustment) |  |
| connecting patients with relevant social care resources (assistance) |  |
| organizing social care assets in the community to improve health outcomes (alignment) |  |
| working with partner social care organizations to promote policies that improve health outcomes (advocacy) |  |

3. In general, is a polysocial risk score more suited for:
   a. Individual-level care activities (e.g. screening, referrals, clinical decision making)
   b. Population-level activities (e.g. risk stratification, quality reporting, risk adjustment)
   c. Both equally
